# A comparison of long-term mortality associated with pathologic placental separation: highlighting possible trends and mechanisms

**DOI:** 10.1101/2025.11.26.25341112

**Authors:** Sona Jasani, Atalay Demiray, Julia Stevenson, Conrad Krawiec

## Abstract

Abruption and retention, two types of abnormal placental separation, are associated with significant morbidity and mortality. Though advancements in obstetric management have improved peripartum injury, patients who experience abnormal placental separation may be at risk for long-term complications. This study evaluates long-term mortality in patients with abruption or retention compared to those with normal placental separation. When controlling for demographic factors, covariates and short-term mortality events, long-term mortality remained significantly elevated in the retention group and was lost in the abruption group. A variety of health outcomes were associated with either abruption, retention or in both abnormal placental separation groups. More research is needed to understand the mechanisms associated with abnormal placental separation and contributors to long-term mortality.

## INTRODUCTION

Two primary complications of placental separation are abruption and retention. Placental abruption involves the complete or partial separation of a normally implanted placenta before delivery. Placental retention happens when the placenta does not fully separate within 30-60 minutes after birth. Both conditions pose significant risks. Abruption can lead to severe complications such as hemorrhage, need for transfusions, disseminated intravascular coagulopathy (DIC), renal failure, and even mortality [1–5]. Similarly, placental retention can result in hemorrhage, invasive procedures, endomyometritis and mortality [6–8].

While advancements in obstetric care have significantly reduced placental separation associated mortality [3,9–11], individuals with placental separation complications may still be at risk for long-term health issues. Research indicates that these two conditions can heighten the risk of cardiovascular, neurological and oncological problems [12–14]. Specifically, patients who experience abruption have an elevated long-term risk of cardiovascular and cerebrovascular diseases [1,12,15–21], while those who experience placental retention accompanied by hemorrhage face increased risks of cardiovascular disease and cancer [13]. These findings highlight the need for ongoing monitoring and preventative care for individuals who have experienced these complications, emphasizing the importance of early recognition and intervention to mitigate long-term health risks.

The reasons behind these associations remain unclear [22–25]. While biological factors likely play a role, the clinical approach to post-delivery management of patients who experience abnormal placental separation may also contribute. The absence of standardized protocols for postpartum follow-up may lead to missed or delayed diagnoses and/or treatment, disproportionately affecting some populations. This uncertainty is also complicated by limited mechanistic understanding of the etiology and outcomes of placental dysfunction. The tendency to examine abnormal placental separation into their separate clinical diagnoses, abruption or retention, may impede the identification of molecular pathways that underly placental dysfunction perinatally and beyond. Placental dysfunction can include conditions such as abruption, preeclampsia, fetal growth restriction or preterm delivery and are associated with long-term cardiovascular diseases [25–26]. Maternal vascular malperfusion has been found in all these conditions [26] and may be one etiologic component of placental dysfunction. Though retained placenta is not traditionally categorized under placental dysfunction, similarities such as maternal vascular malperfusion [27–28] and associated long-term outcomes [13–14] suggests mechanistic similarities. Categorizing abruption and retention under the broader concept of abnormal placental separation may improve our understanding of the pathogenesis and inform both acute and long-term care strategies.

The primary objective of this study is to compare long-term mortality in patients with pathologic placental separation in an index delivery. We hypothesize that patients with either type of abnormal placental separation, abruption or retention, will have a higher risk of long-term mortality as compared to normally separating placentas. We chose to focus on a clear outcome such as mortality as this is clinically significant and accurately represented in electronic health records (EHR). The secondary objective of this study is to analyze associations between abruption and retention with health outcomes to further elucidate potential disease mechanisms and risk factors.

## MATERIALS AND METHODS

### Study design

A retrospective observational case control cohort study was conducted using the TriNetX EHR database of all women who delivered vaginally between the ages of 15-54 with a reported inpatient encounter. TriNetX is a global federal health research network that provides researchers access to continuously updated data elements from participating health care organizations, predominantly in the United States. TriNetX is certified to the ISO 27001:2013 standard and protects healthcare data by maintaining compliance with the Health Insurance Portability and Accountability Act (HIPPA) Security Rule. The EHR data elements are aggregated, de-identified, and include demographic characteristics, diagnoses, procedures, medications, laboratory values and genomics, all in compliance with the de-identification standard outlined in Section 164.51 (a) of the HIPPA privacy rule. As no protected health information is received by the user, we were provided a waiver from the Penn State Health Institutional Review Board to perform this study (STUDY00020794). Study design, conduct and result reporting were constructed using the Strengthening the Reporting of Observational Studies in Epidemiology (STROBE) guidelines.

### Data collection

All deliveries were identified in TriNetX on April 5, 2024, using related common procedural terminology (CPT) codes for vaginal deliveries and cesarean section (N=1,483,438). The earliest delivery date was identified for all patients and used to determine mode of delivery (i.e. vaginal or cesarean section). We then identified those deliveries with a placental separation code using the specific International Classification of Diseases (ICD) codes for placental abruption and retained placenta (data in S1 Appendix). Dates for placental separation ICD codes were identified in order to ensure that placental separation status co-occurred with the identified delivery for each patient. Placental separation ICD codes 6 weeks before delivery and up to 6 weeks after delivery were used to assign a delivery to a placental outcome group. Deliveries that did not have co-occurring placental separation ICD codes were excluded in the primary analysis but were retained for the purposes of sensitivity analysis for mortality (data in S2 Appendix).

### Patients without any ICD codes for placental separation were assigned to the normal delivery group

We then extracted the earliest and latest encounter dates for each patient. Patients with less than 30 days of follow-up, with records indicating both abruption and retention, and those delivered by cesarean section were excluded to ensure mutually exclusive groupings and to control for the possible impact of delivery mode on maternal morbidity and mortality. The final dataset comprised of 638,911 patients after applying these inclusion and exclusion criteria. We then obtained the following data: age, race, ethnicity, Centers for Disease Control and Prevention (CDC) classification-related maternal morbidity diagnoses, CPT codes, healthcare common procedure coding system (HCPS) codes, and relevant health outcomes using ICD codes [29–30]. In order to control for possible differences in medical record data entry, we excluded deliveries prior to January 1, 2008, as EHR use prior to 2008 was inconsistent (Fig 1). The primary outcomes assessed were mortality within 365 days of delivery and time to death within specified intervals, including the full follow-up period (up to 3000 days), mortality within 365 days (censoring events beyond this period), and mortality between 30 and 365 days (excluding deaths within the first 30 days).

**Fig 1.**
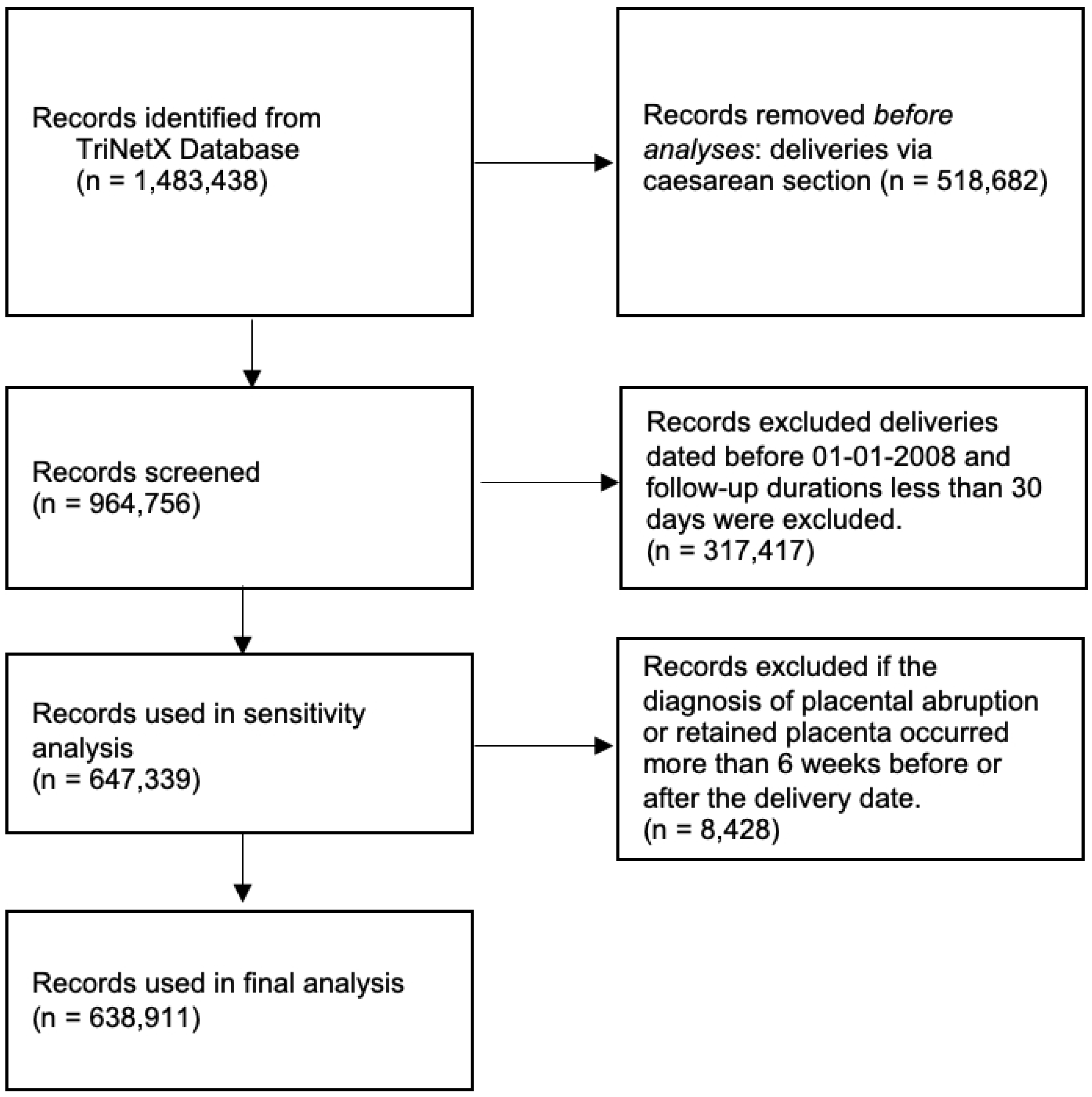
Data Collection Flowchart

### Statistical analysis

Categorical variables, including demographic factors and health conditions, were summarized as counts and percentages. Comparisons between groups were performed using the Chi-square test. Continuous variables, such as maternal year of birth and follow-up duration, were summarized as means and standard deviations. Since continuous variables were not normally distributed (assessed visually via histograms and confirmed by Shapiro-Wilk tests), comparisons across groups were performed using the Kruskal-Wallis test, a non-parametric method appropriate for non-normally distributed data.

Cox proportional hazards regression models were used to estimate hazard ratios (HRs) and 95% confidence intervals (CIs) for the association between placental outcomes and subsequent mortality. Both unadjusted and adjusted models were fitted (data in S3 Appendix). Adjusted models controlled for potential confounders, including maternal age, race/ethnicity, and social determinants of health (SDOH) indicator, and other diagnosed health conditions. For each Cox proportional hazards model, the proportional hazards assumption was evaluated both graphically using Schoenfeld residuals and formally tested using the global test. Models where the global test p-value was less than 0.05 were considered to violate the proportional hazards assumption.

Kaplan-Meier survival curves were generated to visualize survival probabilities across groups, and differences in survival distributions were formally tested using the log-rank test. For time-specific mortality outcomes, such as deaths occurring beyond 30 days and 365 days after delivery, survival times were censored accordingly (data in S4 Appendix). In addition to mortality, associations between placental outcomes and a range of diagnosed health conditions (including hypertension, cardiovascular disease, sepsis, among others) were investigated. For each health outcome, univariable Cox regression models were first fit to estimate unadjusted HRs with 95% CIs. Subsequently, multivariable Cox regression models were constructed, adjusting for maternal age, race/ethnicity, and SDOH indicator.

Covariates included in multivariable models were selected based on theoretical relevance and data availability. No stepwise selection methods were applied. Multicollinearity among covariates was assessed using variance inflation factors (VIFs), and no VIF exceeded the threshold of 5.

HRs with 95% CIs and corresponding p-values are reported. All hypothesis tests were two-sided, and a p-value < 0.05 was considered statistically significant. No corrections for multiple comparisons were applied due to the exploratory nature of secondary analyses, though findings are interpreted cautiously. All statistical analyses were conducted using R software, version 4.3.1 (R Foundation for Statistical Computing, Vienna, Austria). The complete R code used to perform the analyses is available (data in S5 Appendix). We cannot share the raw data file because it is licensed by TriNetX and based on their policies they do not allow sharing.

## RESULTS

The majority of deliveries were categorized into the normal placental separation group (97.96%). Approximately 1.19% of deliveries were categorized into the retention group and 0.85% were categorized into the abruption group, consistent with the reported prevalence of these conditions respectively. Significant demographic differences were noted between normal and abnormal placental separation groups (Table 1). Additionally, significant differences were also identified in a majority of the health outcomes analyzed in our data set. Conditions that did not significantly differ included breast cancer, colorectal cancer, cystic fibrosis and HIV.

**Table 1.**
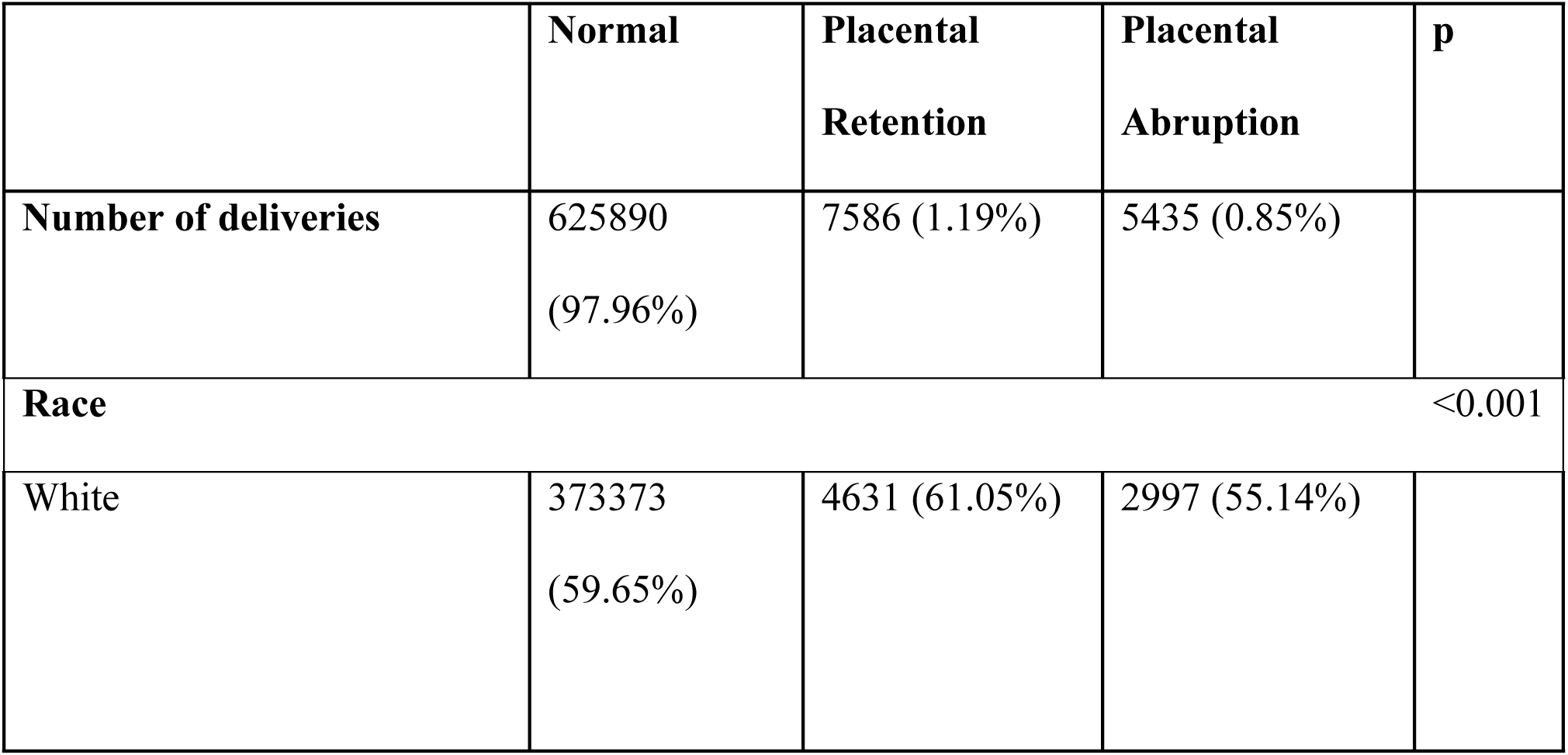
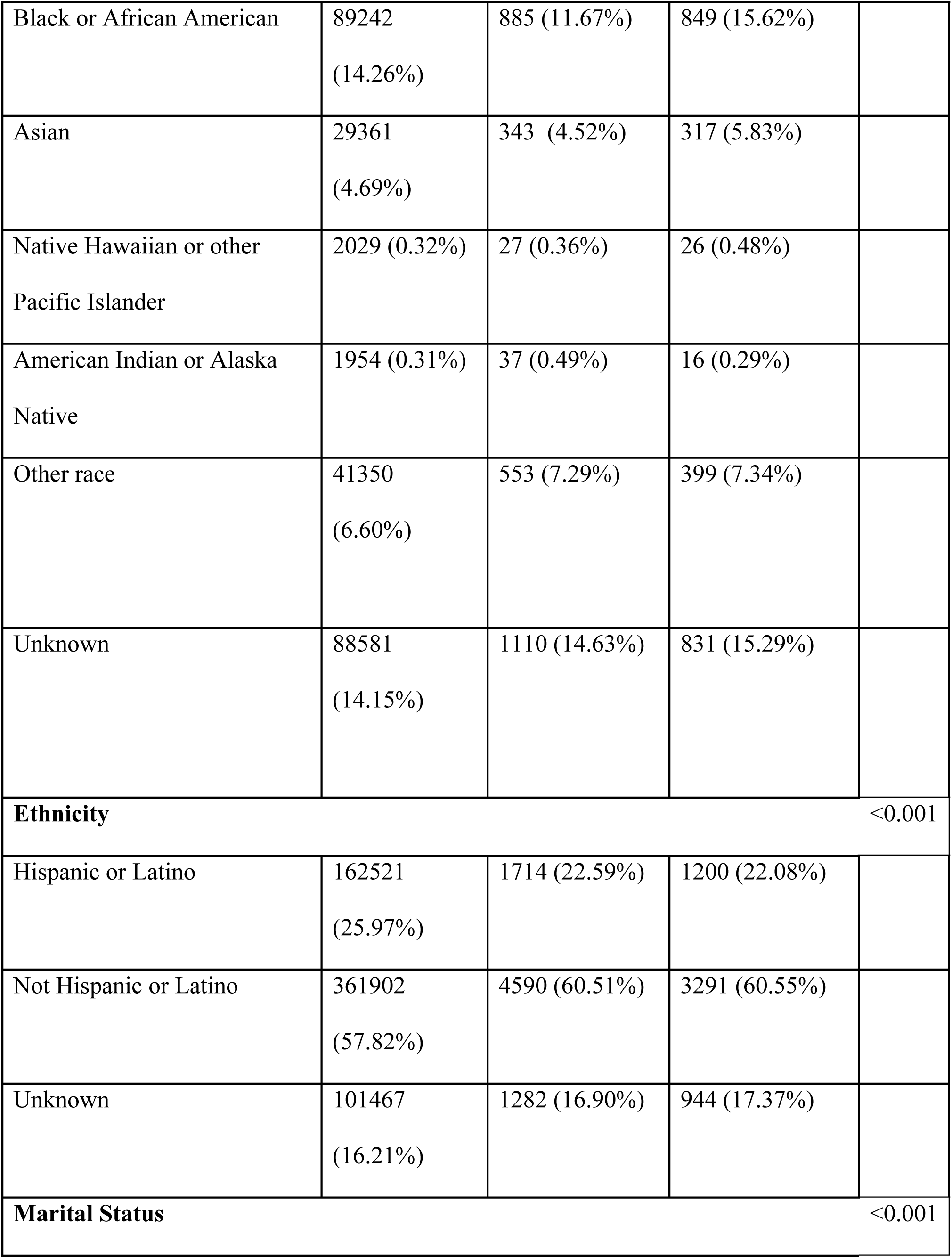
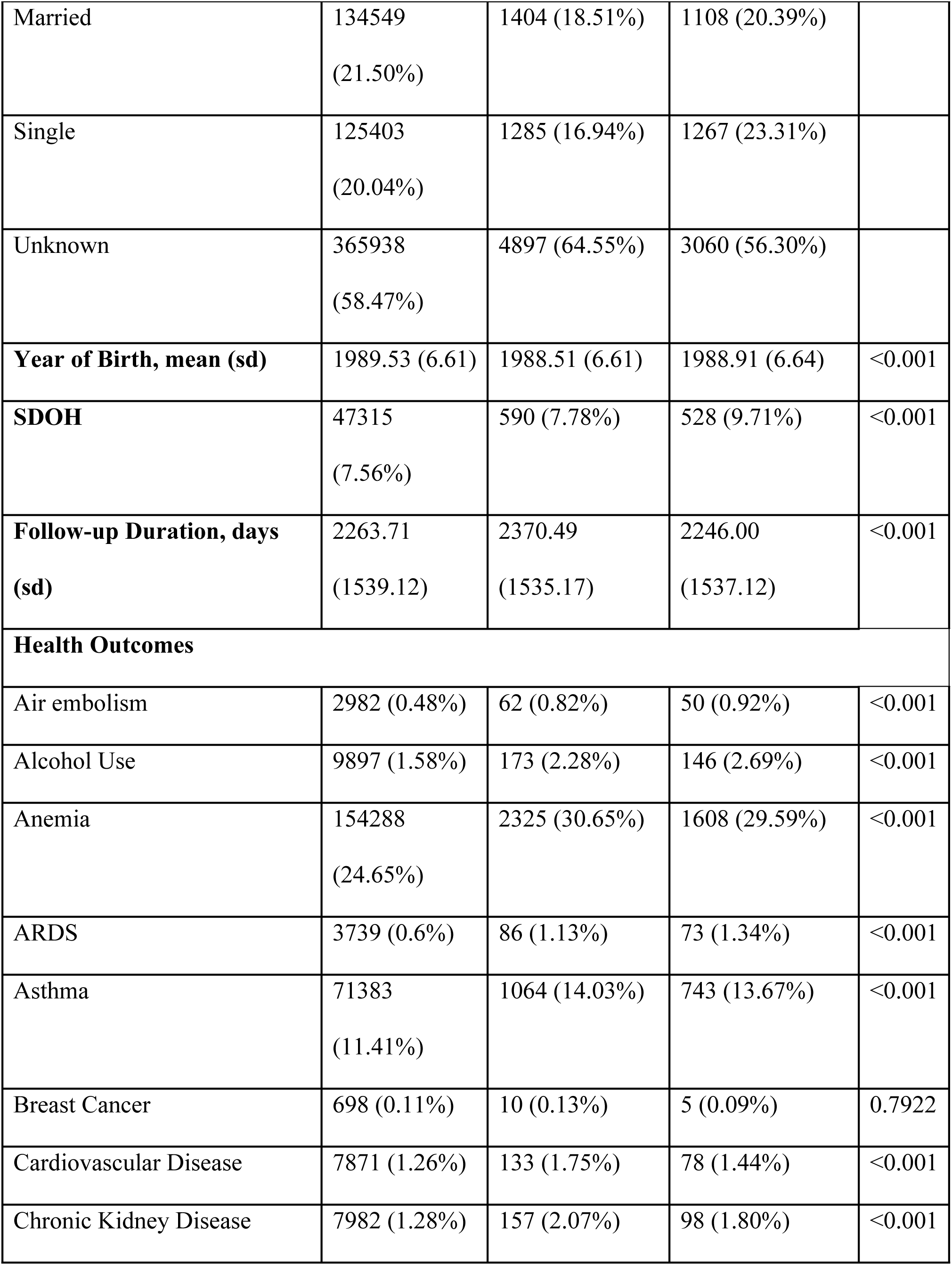
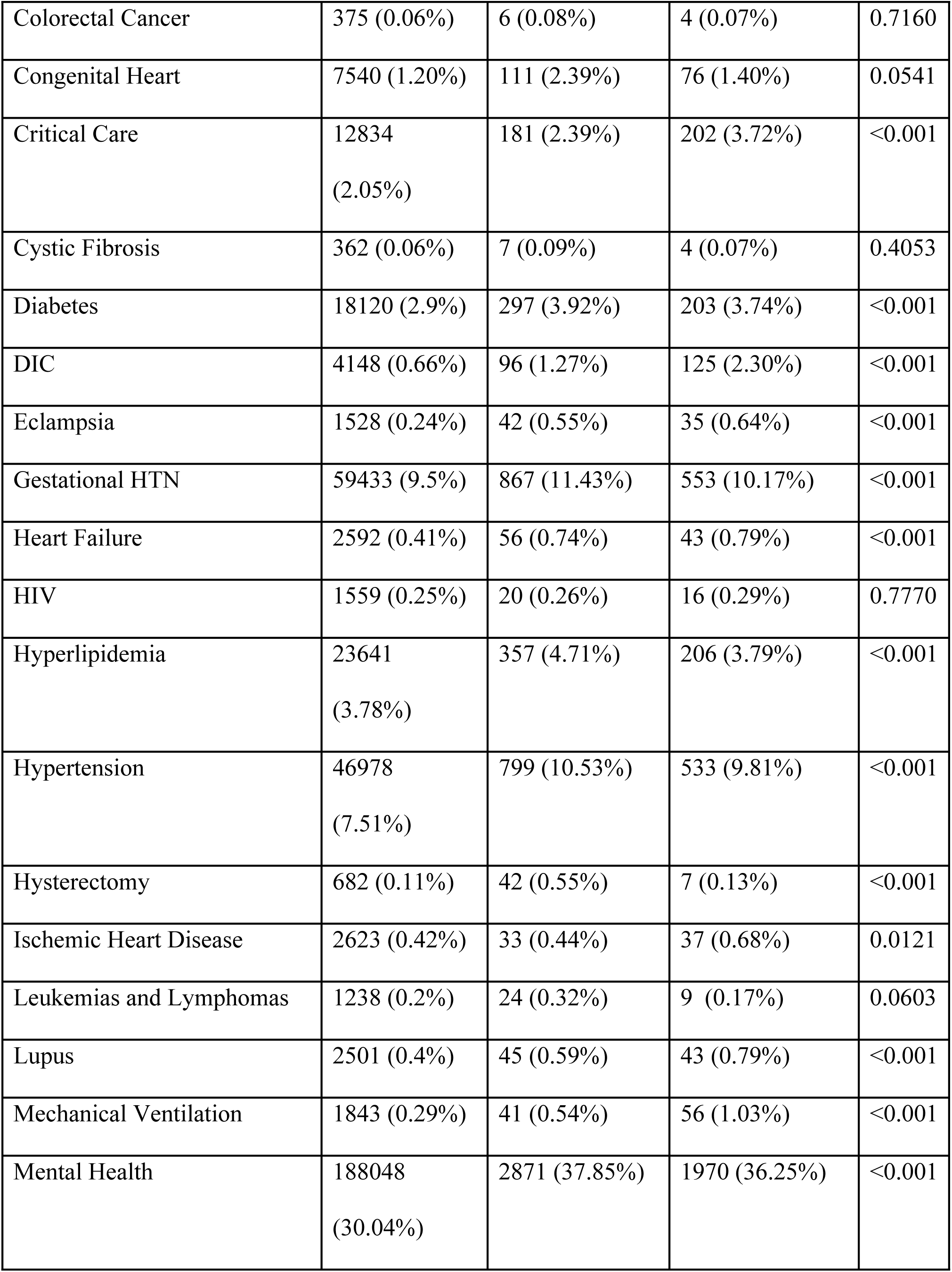
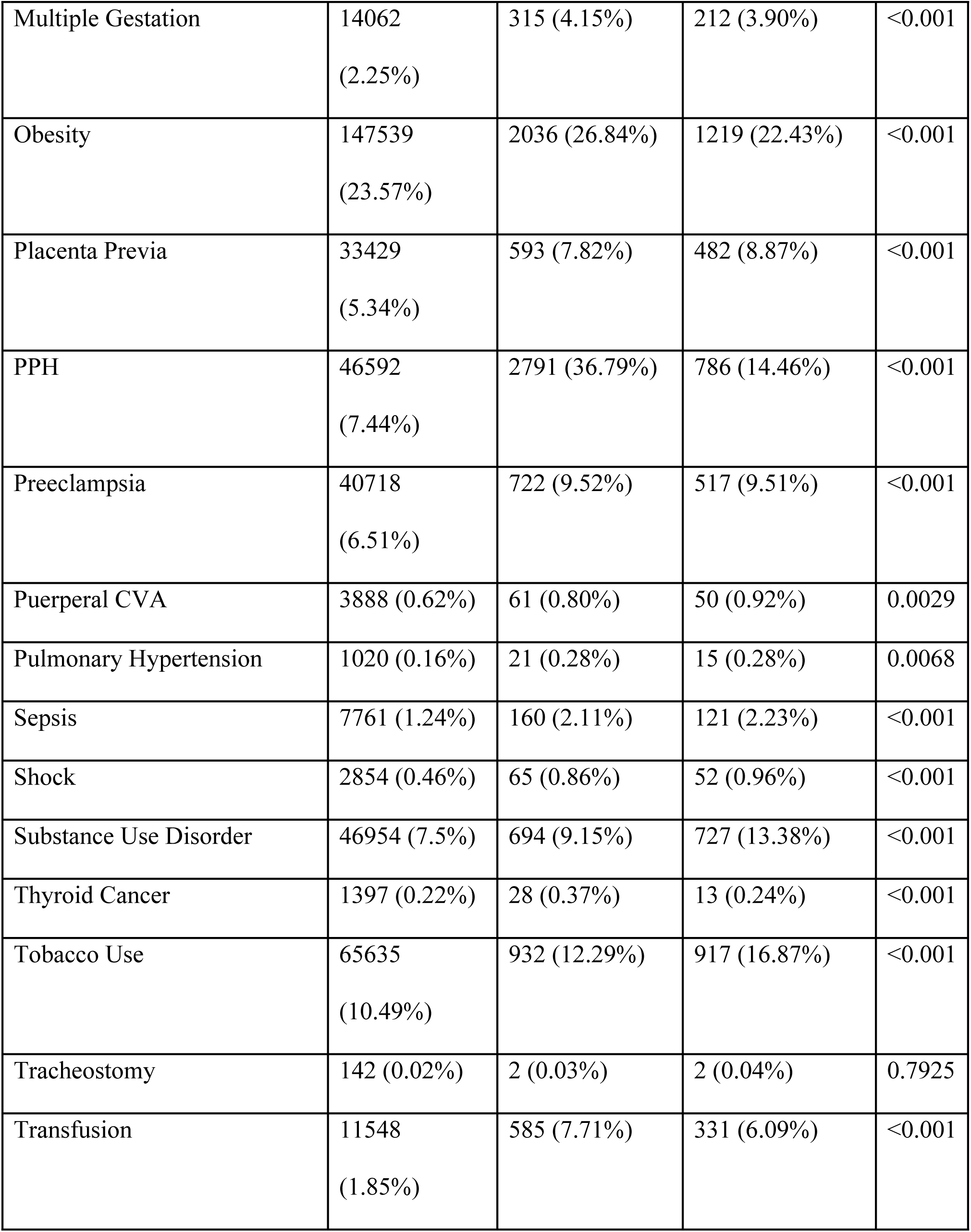
Demographic factors for patients with abruption, retention and normal placental separation.

Mortality HR (Table 2) for patients who experienced either abruption or retention were statistically significantly elevated in unadjusted calculations and when adjusted for demographic factors (age, race, SDOH). When adjusted for demographic factors and all health outcomes accounted for in our study, only the mortality HR for retained placenta remained significantly elevated. Evaluation of the proportional hazards assumption revealed no violation in the unadjusted mortality model (global p = 0.81); however, violations were observed in adjusted models (global p = 0.011) To address potential time-dependent effects, particularly those arising from acute postpartum mortality, we conducted sensitivity analyses excluding deaths occurring within the first 30 days and 365 days following delivery. When excluding deaths within 365 days post-delivery (thus evaluating mortality among longer-term survivors), the proportional hazards assumption for the fully adjusted mortality model was no longer violated (global p = 0.069).

**Table 2.** Unadjusted and adjusted hazard ratio of mortality associated with pathologic placental separation of all patients, of those who survived beyond 30 days of delivery and of those who survived one year beyond delivery.

| **Group** | **HR unadj** | **p** | **HR adj**<br>**(demographic)** | **p** | **HR adj**<br>**(demographic**<br>**+ all outcomes)** | **p** |
| --- | --- | --- | --- | --- | --- | --- |
| **Abruption**<br>**all** | 1.61<br>[1.23,2.12] | 0.0005 | 1.59<br>[1.20,2.08] | 0.0009 | 1.16<br>[0.89,1.53] | 0.277 |
| **Abruption**<br>**– 30 days** | 1.35<br>[0.99,1.83] | 0.0583 | 1.32<br>[0.97,1.80] | 0.0773 | 0.96<br>[0.71,1.31] | 0.8023 |
| **Abruption**<br>**– 1 year** | 1.52<br>[1.11,2.10] | 0.0098 | 1.49<br>[1.08,2.06] | 0.0140 | 1.09<br>[0.79,1.50] | 0.6173 |
| **Retention**<br>**all** | 2.04<br>[1.66,2.51] | <0.0001 | 1.95<br>[1.59,2.40] | <0.0001 | 1.48<br>[1.20,1.83] | <0.0001 |
| **Retention -**<br>**30 days** | 2.03<br>[1.63,2.52] | <0.0001 | 1.94<br>[1.56,2.41] | <0.0001 | 1.48<br>[1.19,1.84] | 0.0005 |
| **Retention**<br>**– 1 year** | 2.12<br>[1.67,2.68] | <0.0001 | 2.02 [1.59,2.55] | <0.0001 | 1.55<br>[1.22,1.97] | 0.0003 |

Importantly, the mortality HR for retained placenta remained significantly elevated across all models, including those excluding deaths within 30 and 365 days. In contrast, for placental abruption, the initially observed mortality risk lost statistical significance after excluding early postpartum deaths (i.e., deaths within 30 days), suggesting that elevated mortality risk associated with abruption was primarily driven by short-term mortality in the immediate postpartum period.

Health outcomes analyzed included CDC-related maternal morbidity diagnoses, related CPT, HCPS codes, and predefined ICD codes [29–30]. Conditions that were statistically significantly associated with either abruption, retention or both after adjusting for demographic factors (P value <0.05) were identified and organized into the following categories: abnormal placentation, obstetric associations, health behaviors, health system, chronic conditions, acute conditions, or either chronic or acute conditions (Table 3). Conditions that were significantly increased in both abruption and retention included: placenta previa, preeclampsia, eclampsia, gestational hypertension, multiple gestation, PPH, transfusion, mechanical ventilation, critical care use, chronic hypertension, diabetes, chronic kidney disease, mental health, lupus, asthma, anemia, acute respiratory distress syndrome, air embolism, shock, disseminated intravascular coagulopathy and heart failure. Conditions that were significantly elevated in abruption only included: congenital heart disease, ischemic heart disease, and puerperal cerebrovascular disease. Conditions that were significantly elevated in retention only included: cardiovascular disease, hysterectomy, pulmonary hypertension and thyroid cancer. Obesity was the only condition that was significantly associated with both abruption and retention that had differing directions of risk association (decreased in abruption [HR 0.93] and increased in retention [HR 1.12]).

**Table 3.** Significant clinical associations with pathologic placental separation states after adjusting for demographic factors.

| CATEGORY | Placental<br>Retention<br>(adj HR) | Placental<br>Abruption<br>(adj HR) | Comparative<br>Directionality |
| --- | --- | --- | --- |
| **Abnormal placentation** |  |  |  |
| Placenta previa | 1.36 [1.25, 1.47]<br>p<0.001 | 1.69 [1.54, 1.85]<br>p<0.001 | Similar |
| **Obstetric** |  |  |  |
| Preeclampsia | 1.45 [1.35, 1.56]<br>p<0.001 | 1.49 [1.37, 1.63]<br>p<0.001 | Similar |
| Eclampsia | 2.24 [1.64, 3.04]<br>p<0.001 | 2.57 [1.84, 3.59]<br>p<0.001 | Similar |
| Gestational hypertension | 1.16 [1.09, 1.25]<br>p<0.001 | 1.08 [0.99, 1.18]<br>p<0.007 | Similar |
| Multiple gestation | 1.77 [1.58, 1.98]<br>p<0.001 | 1.73 [1.51, 1.98]<br>p<0.001 | Similar |
| PPH | 5.46 [5.25, 5.67]<br>p<0.001 | 2.01 [1.88, 2.17]<br>p<0.001 | Similar |
| **Health behaviors** |  |  |  |
| Alcohol | 1.45 [1.25, 1.68]<br>p<0.001 | 1.70 [1.45, 2.00]<br>p<0.001 | Similar |
| Tobacco | 1.22 [1.15, 1.30]<br>p<0.001 | 1.72 [1.61, 1.84]<br>p<0.001 | Similar |
| Substance use | 1.28 [1.18, 1.38]<br>p<0.001 | 1.88 [1.74, 2.02]<br>p<0.001 | Similar |
| **Health System** |  |  |  |
| Transfusion | 4.26 [3.92, 4.62]<br>p<0.001 | 3.35 [3.00, 3.74]<br>p<0.001 | Similar |
| Mechanical ventilation | 1.76 [1.29, 2.39]<br>p<0.001 | 3.54 [2.71, 4.62]<br>p<0.001 | Similar |
| Critical care use | 1.21 [1.04, 1.40]<br>p<0.012 | 1.89 [1.65, 2.17]<br>p<0.001 | Similar |
| Hysterectomy | 4.15 [3.04, 5.67]<br>p<0.001 | 1.12 [0.53, 2.36]<br>p=0.765 | Different<br>(sig. R only) |
| **Chronic** |  |  |  |
| Hypertension | 1.29 [1.20, 1.38]<br>p<0.001 | 1.27 [1.16, 1.38]<br>p<0.001 | Similar |
| Diabetes | 1.20 [1.07, 1.35]<br>p=0.001 | 1.19 [1.04, 1.37]<br>p=0.012 | Similar |
| Chronic kidney disease | 1.57 [1.34, 1.84]<br>p<0.001 | 1.42 [1.16, 1.73]<br>p<0.001 | Similar |
| Mental health | 1.27 [1.22, 1.31]<br>p<0.001 | 1.29 [1.23, 1.35]<br>p<0.001 | Similar |
| Lupus | 1.38 [1.03, 1.86]<br>p=0.031 | 1.92 [1.42, 2.60]<br>p<0.001 | Similar |
| Asthma | 1.25 [1.17, 1.33]<br>p<0.001 | 1.22 [1.13, 1.31]<br>p<0.001 | Similar |
| Cardiovascular disease | 1.27 [1.07, 1.51]<br>p<0.001 | 1.13 [0.91, 1.41]<br>p=0.278 | Different<br>(sig. R only) |
| Congenital heart disease | 1.16 [0.97, 1.40]<br>p=0.112 | 1.17 [0.94, 1.47]<br>p=0.167 | Similar |
| Pulmonary hypertension | 1.55 [1.01, 2.39]<br>p=0.046 | 1.56 [0.94, 2.60]<br>p=0.087 | Different<br>(sig. R only) |
| Thyroid cancer | 1.40 [0.96, 2.03]<br>p=0.007 | 1.03 [0.60, 1.79]<br>p=0.901 | Different<br>(sig. R only) |
| Obesity* | 1.12 [1.07, 1.17]<br>p<0.001 | 0.93 [0.88, 0.98]<br>p=0.012 | Different<br>(sig. R&A) |
| **Acute** |  |  |  |
| Acute respiratory distress syndrome | 1.85 [1.49, 2.29]<br>p<0.001 | 2.21 [1.75, 2.78]<br>p<0.001 | Similar |
| Air embolism | 1.60 [1.24, 2.05]<br>p<0.001 | 1.87 [1.41, 2.47]<br>p<0.001 | Similar |
| Shock | 1.81 [1.42, 2.32]<br>p<0.001 | 2.08 [1.58, 2.73]<br>p<0.001 | Similar |
| Sepsis | 1.70 [1.46, 1.99]<br>p<0.001 | 1.80 [1.50, 2.16]<br>p<0.001 | Similar |
| Puerperal cerebrovascular disease | 1.18 [0.91, 1.52]<br>p=0.205 | 1.44 [1.09, 1.90]<br>p=0.0107 | Different<br>(sig. A only) |
| **Either acute or chronic** |  |  |  |
| Disseminated intravascular coagulopathy | 1.79 [1.47, 2.20]<br>p<0.001 | 3.41 [2.86, 4.08]<br>p<0.001 | Similar |
| Heart failure | 1.63 [1.25, 2.13]<br>p<0.001 | 1.79 [1.33, 2.42]<br>p<0.001 | Similar |
| Anemia | 1.28 [1.24, 1.34]<br>p<0.001 | 1.21 [1.16, 1.27]<br>p<0.001 | Similar |
| Ischemic heart disease | 0.91 [0.65, 1.29]<br>p=0.6 | 1.53 [1.11, 2.12]<br>p=0.009 | Different<br>(sig. A only) |

R=Retention, A=Abruption
All conditions in the similar column were significantly elevated in both abruption and retention groups after adjustment. All conditions in the different column were significantly elevated in either abruption or retention groups with the exception of obesity.
\*Obesity was significantly elevated in retention and significantly reduced in abruption groups respectively.

## DISCUSSION

Our primary aim was to compare long-term mortality in those with pathologic placental separation to those with normal separation. Both abruption and retention were associated with significantly increased mortality in unadjusted models and when adjusted for demographic factors. However, after further adjustment for comorbid conditions, the association with abruption was no longer significant. Excluding deaths within the first 365 days postpartum did not change mortality risk for either group but excluding deaths within 30 days attenuated the association for abruption while the elevated risk for retention persisted across all models. These findings highlight the importance of distinguishing between types of abnormal placental separation when assessing long-term prognosis and suggest that women with retained placenta may benefit from targeted follow-up and preventive care after delivery. The distinct risk patterns we observed raise important questions about underlying mechanisms and potential intervention points that warrant further investigation.

Abruption occurs when the placenta separates prior to neonatal delivery and retention occurs when the placenta does not separate after neonatal delivery. Both abruption and retention are associated with serious obstetric hemorrhage resulting in acute maternal morbidity and mortality [1–8,15,31–36]. Abruption-associated mortality is seven times higher than overall maternal mortality [3] and retention associated mortality can reach up to 10% globally [8]. In the U.S., abruption associated mortality ranges from 1-5% [2] and in-hospital mortality from retention is approximately 2.8% [7]. Consistent with the literature, our results show significantly increased acute health interventions such as critical care use, transfusion, and mechanical ventilation in both abruption and retention groups. Knowledge gaps however remain in understanding the long-term implications of surviving abnormal placental separation. Abruption is associated with long-term maternal mortality and morbidity from both cardiovascular and non-cardiovascular etiologies [15–21]. Similarly, retention with hemorrhage is associated with long-term cardiovascular and cancer risks [13]. Though placental dysfunction and cardiovascular disease have mechanistic similarities [1,12,32,37], how these mechanisms contribute to long-term morbidity and mortality is currently unknown. Answering these questions is clinically relevant in guiding both postpartum care and the obstetric to primary care transition. Presently, postpartum follow-up recommendations do not differ between patients with abnormal placental separation from those with normally separating placentas. Our data supports close postpartum monitoring for vascular or hematologic dysfunction in those with abruption as well as continued monitoring for cardiovascular disease after transitioning out of obstetric care in those with retention.

Our results highlight the importance of understanding abnormal placental separation mortality trends. We found that long-term mortality is affected by short-term mortality events (deaths within 30 days of delivery) in those with abruption but not in those with retention.

Maternal mortality reviews typically organize etiologies into broad categories (i.e. obstetric hemorrhage) that encompass a variety of pathologies (atony, abruption, retention, previa, uterine rupture) [38–41] with a focus on analyzing outcomes during pregnancy and within one year of delivery [39–40]. This broad approach however can obfuscate the mortality trends of each individual pathology. By comparing abruption and retention, our study adds to the existing literature on this topic. In the abruption group only, we found significantly increased prevalence of congenital heart disease, ischemic heart disease and puerperal cerebrovascular accidents. All of these conditions are associated with maternal mortality during delivery or postpartum [42–45] and may explain why elevated mortality HR significance was lost in the abruption group when we adjusted for all health outcomes and controlled for short-term mortality. In the retention only group, we found significantly increased prevalence of cardiovascular disease and thyroid cancer. These conditions may be more insidious as compared to the conditions of the abruption only group and may not have impacted long-term mortality in the same way as was seen with abruption. In contrast to the literature [13], hemorrhage did not explain the increased long-term mortality in the retention group because we included this health outcome as a covariate in adjustments. It is unclear if the nature of retention itself may be contributing to this elevated long-term mortality risk. Our differences may be due to small numbers, a shorter mean follow-up duration (7.2 years ± 4.1) or the exclusion of deliveries by cesarean section. Identifying the dysregulated pathways in retention that contribute to long-term mortality is necessary to mitigate these risks.

The secondary aim of this study was to explore possible disease mechanisms associated with abnormal placental separation. By comparing abruption and retention groups, we found a significantly increased prevalence of placenta previa and preeclampsia/eclampsia in abruption and retention. Mechanisms contributing to implantation, placental migration, matrix metalloproteinases, apoptosis, inflammatory cytokine expression and uterine/endometrial blood supply may explain the association with placenta previa [33–34,46–47]. Angiogenesis related biomarkers such as soluble fms-like tyrosine kinase-a (sFlt-1), placental growth factor (PlGF), and the sFlt-1/PlGF ratio may explain the association with preeclampsia/eclampsia [31,48–51].

The exact molecular mechanisms that underly abruption and retention are currently unknown, but the similarities in proposed mechanisms are striking. Abruption is thought to involve placental hypoperfusion, inadequate placentation, and acute mechanical shearing forces [1]. Similarly, retained placenta is thought to involve placental hypoperfusion, abnormal implantation, and inadequate contractile forces [6,8,52]. The significant similarities between associated health outcomes, abruption and retention demonstrated in our study lends credence to the notion of possible shared mechanisms in pathophysiology. Though our results cannot identify specific biologic mechanisms, inflammation may underly the associations we noted between abnormal placental separation [27–28,53–54] and the variety of health outcomes examined in our study [55–61]. Elucidating the interaction between immune function and abnormal placental separation is therefore a promising area to explore for future studies.

The strengths of this study include our large multi-center study cohort, case control study design, comprehensive health outcome list, and our comparative approach analyzing both abruption and retention. We focused on the clear outcome of mortality and excluded deliveries prior to 2008 to control for possible EHR data quality issues. Furthermore, we adjusted all mortality HR with demographic and health outcomes to control for all differences seen between groups. There are several limitations to this study. Due to database limitations, diagnostic codes were used to categorize placental separation groups and identify all health conditions. Thus, our study may be subject to misclassification bias. Statistically, this misclassification may increase a type II error and possibly miss some significant associations. Furthermore, our study design does not allow for causational or chronologic associations. The elevated health outcomes associated with abnormal placental separation could reflect associations either prior to or following the index delivery. Thus, they should be considered to demonstrate relationships with possible underlying mechanisms that need to be explored in future studies. Finally, we could not fully account for the role that obesity may have played. Our results showed obesity to be elevated in retention and reduced in abruption consistent with the literature [36,48,52,62–63], however, we could not fully control for this covariate due to inconsistent information on BMI or pregnancy weight gain in the TriNetX dataset.

## CONCLUSION

Long-term mortality risk in patients who experience abruption or retention during an index delivery is elevated when adjusting for demographic factors. When adjusting for demographic and all health outcomes analyzed in this study, long-term mortality remained significantly elevated only in those with retention. Short-term mortality may affect long-term mortality in patients with abruption. Comparing and contrasting significant health associations between abruption and retention groups can provide exploratory information to guide further analyses. More research is needed to identify the mechanistic contributors to long-term mortality in those with abnormal placental separation.

## Data Availability

The data that support the findings of this study originate from the TriNetX global federated research network (https://trinetx.com). Because these data are governed by institutional licensing restrictions, we are legally prohibited from sharing them publicly. Per the terms of our data use agreement, TriNetX data cannot be released, posted, or redistributed by study authors. Data access is available to qualified researchers upon request directly to TriNetX at, subject to a separate data use agreement and potential costs. TriNetX will provide access to replicate the analyses for researchers who meet their criteria for data access.

## ACKNOWLEDGEMENTS

none

## SUPPORTING INFORMATION CAPTIONS

**S1 Appendix** Table of placental outcome ICD codes

**S2 Appendix** Table of mortality HR prior to excluding co-occurring abnormal placental separation codes

**S3 Appendix** Figure of hazard ratios for mortality by placental condition

**S4 Appendix** Figure of Kaplan-Meier survival curves following delivery with different placental conditions

**S5 Appendix** R code for statistical analysis

